# Slight increase in fomite route transmission risk of SARS-CoV-2 Omicron variant compared with the ancestral strain in households

**DOI:** 10.1101/2022.04.13.22273821

**Authors:** Shuyi Ji, Shenglan Xiao, Huaibin Wang, Hao Lei

## Abstract

The Omicron SARS-CoV-2 variant has become the dominant lineage worldwide, and experimental study had shown that SARS-CoV-2 Omicron variant was more stable on various environmental surfaces than ancestral strain. However, how the changes of stability on surfaces would influence the role of fomite route in SARS-CoV-2 transmission is still unknown. In this study, we modeled the Omicron and ancestral strain SARS-CoV-2 transmission within a household over 1-day period from multiple pathways, i.e., airborne, droplet and contact route. We assumed there were 2 adults and 1 child in the household, and one of the adults was infected with SARS-CoV-2. We assume a scenario of pre-/asymptomatic infection, i.e., SARS-CoV-2 was emitted by breathing and talking, and symptomatic infection, i.e., SARS-CoV-2 was emitted by breathing, talking, and coughing. In pre-/asymptomatic infection, all three routes contributed a role, contact route contribute most (37%-45%), followed by airborne route (34%-38%) and droplet route (21%-28%). In symptomatic infection, droplet route was the dominant pathway (48%-71%), followed by contact route (25%-42%), airborne route played a negligible role (<10%). In the contact route, indirect contact (fomite) route dominated (contributed more than 97%). Compared with ancestral strain, though the contribution of contact route increased in Omicron variant transmission, the increase was slight, from 25%-41% to 30%-45%.

## Introduction

Since the first emergency of severe acute respiratory syndrome coronavirus 2 (SARS-CoV-2) in December 2019, the virus has spread across the globe and has caused over 469 million cases and 6.1 million deaths as of 21 March 2022 (WHO, 2022). As a respiratory infection, SARS-CoV-2 is believed to be mainly transmitted by direct contact, droplet, fomites, and airborne route (WHO, 2020). Exploring the relative importance of different transmission route is crucial for developing targeted infection control strategies. However, it remains unclear to what extent the different transmission routes contributing to SARS-CoV-2 transmission within human beings.

Modelling studies have evaluated the relative importance of different routes for SARS-CoV-2 wild strain in health-care settings (Jones, 2020; Mizukoshi et al., 2021) or households (Lei et al., 2021), and suggested that droplet and airborne transmission routes predominate over the contact route. Fomites as a mode of SARS-CoV-2 transmission were thought to play an important role at the beginning of the pandemic, with laboratory studies revealing the SARS-CoV-2 could persist on plastic, stainless steel and other surface for hours to days (Chin et al., 2020; Pastorino et al., 2020; Marcel et al., 2020). By May 2020, the World Health Organization (WHO) and other agencies were recommending carefully and thoroughly wash their hands and disinfect those frequently touched surfaces. The importance of fomite in SARS-CoV-2 transmission was first contested in July 2020 (Aboubakr et al., 2021; Katona et al., 2022), with literature strengthening this argument in different settings. The Centers for Disease Control and Prevention (CDC) had stated at the time that “People can be infected with SARS-CoV-2 through contact with surfaces. However, based on available epidemiological data and studies of environmental transmission factors, surface transmission is not the main route by which SARS-CoV-2 spreads, and the risk is considered to be low.” (CDC, 2020).

The newly emerged Omicron SARS-CoV-2 variant was firstly identified on 19 November 2021 in South Africa (Viana et al., 2022), and soon become the dominant lineage worldwide, suggesting its high transmissibility in humans. A recent structural study indicates its spike protein is more stable than the ancestral strain (Zeng et al., 2021), and an experimental study had also shown that SARS-CoV-2 Omicron variant was more stable on various environmental surfaces (Chin et al., 2022). And the transmission of SARS-CoV-2 Omicron via packaging had been reported during the past few months. However, how the changes of stability on surfaces would influence the dominant transmission route of SARS-CoV-2 Omicron is still unknown.

The relative importance of different transmission routes certainly varied under different scenarios (Gao et al., 2021). In this study, we considered a household environment since households are high risk settings for transmission of COVID-19 and are an important factor in wider community spread (Haroon et al., 2020; Harris et al., 2021). It had been reported that most clustered COVID-19 infections in the first wave in China were within families (WHO, 2020; Special Expert Group for Control of the Epidemic of Novel Coronavirus Pneumonia of the Chinese Preventive Medicine Association, 2020), suggesting high rates of intra-family transmission and urged prioritization of studies on risk factors for household transmission.

## Methods

### Environmental setting

Given that the mean household size is 2.95 in China (China Statistical Yearbook 2019), and 2.52 in USA. Thus the household size was assumed to be 3, i.e. two parents and one child with age around 10-year-old. Since adults are more susceptible to SARS-CoV-2, and there is no gender difference in susceptibility to COVID-19 (WHO, 2020), thus one of the parents was assumed to be infected with SARS-CoV-2, and other two individuals were assumed to be susceptible to SARS-CoV-2. In this study, we modeled the infection risk of SARS-CoV-2 of two susceptible individuals during 1-day exposure in the household environment.

### Virus shedding by infector

For SARS-CoV-2 infected individuals, they shed viruses into the environment via respiratory activities, such as breathing, talking, coughing, and sneezing. Given that the proportion of asymptomatic/pre-symptomatic infection could reach 57.5% (Yanes-Lane et al., 2020). In this study, we considered the following two scenarios: 1) asymptomatic or presymptomatic infection, i.e. infected individuals shed viruses into the environment via breathing and talking. Breathing and talking for 2 minutes would expel 2.4× 10^−6^ and 7.1× 10^−2^ mL saliva, respectively. During asymptomatic or presymptomatic infection, we assumed that 49.5% and 49.5% of the exhaled droplets by the patient were partitioned to porous and non-porous surfaces, and the rest 1% deposited on the infector’s hands. 2) symptomatic infection, i.e. infected individuals shed viruses into the environment via breathing, talking and coughing. And one cough would expel 0.17 mL saliva, and the cough frequency was 30 per hour. And during symptomatic infection, we assumed that 89% and 10% of the exhaled droplets by the patient were partitioned to porous and non-porous surfaces respectively (Mizukoshi et al., 2021), since the patient would lie in bed most time. The rest 1% deposited on the infector’s hands. Detailed size distribution of droplets from breathing, talking and coughing are in Supplementary Information Part 1. Given that the viral load in the nasopharyngeal swabs peaked, on average, 1 d before symptom onset, with values of 8∼9 log10 RNA copies per mL, and then decreased exponentially (Néant et al., 2021; Wyllie et al., 2020). Thus, in this study, in asymptomatic or presymptomatic infection, the viral concentration in the droplets was set from 10^5^∼10^9^ RNA copies per mL, and in symptomatic infection, the viral concentration was set from 10^4^∼10^8^ RNA copies per mL.

### Transmission route definition

As a respiratory infection, SARS-CoV-2 could be transmitted by contact, droplet, fomites, airborne route, and possible feces-oral, bloodborne and intrauterine transmission (WHO, 2020). In this study, we considered the four major transmission routes, i.e., contact, droplet, fomites, and airborne route, and classified them into the following three categories:

1. The airborne route refers to direct inhalation of an infectious agent through small droplet nuclei, that is, the residue of large droplets containing microorganisms that have evaporated to an aerodynamic diameter of less than 10 microns (termed respirable) (Nicas and Jones, 2009). These respirable droplets can deposit in the respiratory tract.
2. Droplet route refers to the inhalation of the virus carried in respirable airborne particles with a diameter between 10 and 100 microns (termed inspirable) (Nicas and Jones, 2009), and the droplet spray of large droplets (>100 microns in diameter) onto facial target membranes.
3. Contact route includes direct and indirect contact. Direct contact refers to infection transmission through person-to-person body contact, such as handshaking. Indirect contact route is also called fomite route, referring to infection transmission by touching objects or surfaces that have earlier been contaminated by hands or by direct deposition of infectious pathogens from the infected individuals. Direct and indirect contact routes are hard to distinguish since hands could be contaminated via both routes, and the virus concentration on susceptible individuals’ hands sequentially influenced the exposure level. So in this study, we compared the relative importance of direct and indirect contact routes via the infection risk when the susceptible individuals did not touch fomites or hands of the infector.

### The exposure pathway model

A Markov chain was used to model the movement of virus between select compartments in a household environment (Figure 1). We have used the model to study the multi-route infection transmission in airplane, household and hospital (Lei et al., 2018; Xiao et al., 2018; Lei et al., 2021). A total of 8 compartments were considered in this study. Virus is emitted into room air (Compartment 1) in the exhaled droplets by the infector. Some of the virus in the room air could deposit on porous (Compartment 2), non-porous surfaces (Compartment 3), infector’s hands (Compartment 4), exhausted by ventilation (Compartment 5) and inhaled by susceptible individuals (Compartment 6). Virus could transfer between porous/non-porous surfaces and infector’s or susceptible individuals’ hands (Compartment 7) during hand-to-surface contact. The virus on susceptible individuals’ hands could transmitted to the mucus membranes and lead to infection transmission (Compartment 8).

**Figure 1.**
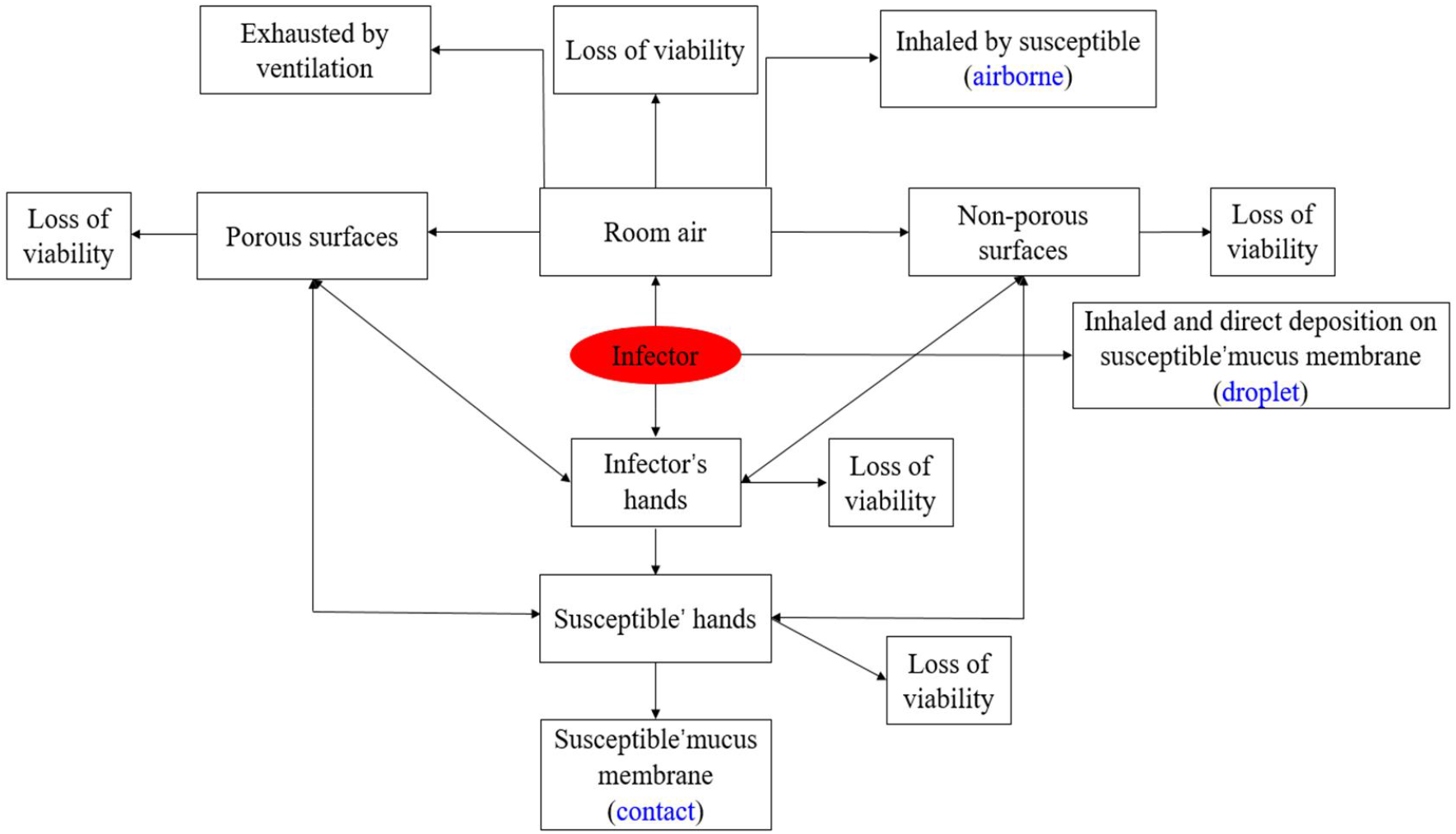
The exposure pathway model in a household environment.

### Airborne route model

Denoted the concentration of droplet nuclei of radius *r* at time t in the room air be *C*_*a*_(*r, t*), *T*_*a*_ is the exposure times of susceptible person in the household, then the exposure dose of susceptible person during *T*_*a*_ exposure time could be calculated by equation (1):

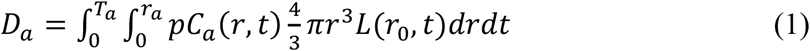

Where *r*_*a*_ is the largest radius for airborne droplets, *p* is the pulmonary ventilation rate. *r*_0_ is the droplets’ initial radius just after exhalation, and we assume that the final radius *r* after complete evaporation is *r* = *r*_0_/3 (Lei et al., 2018). *L*(*r*_0_, *t*) is the concentration of viable virus in droplet with initial radius *r*_0_ after exhalation time t. The concentration of viable pathogens in droplets varies with exhalation time. Studies have shown that pathogens die rapidly in the evaporation process of droplets. For droplets with a radius of less than 5 *μm*, the time from exhalation to evaporation to droplet nuclei is less than 0.1 s (Xie et al., 2006). Therefore, we assumed that after evaporation, only 25% of the virus in the initial droplets was viable in the droplet nuclei (Xie et al., 2006). *C*_*a*_(*r, t*) is the concentration of droplet nuclei of radius *r* in the room air. Without considering virus inhalation by individuals in the household and deposition on vertical environmental surface during the 1-day simulation period because of their negligible contribution, there is

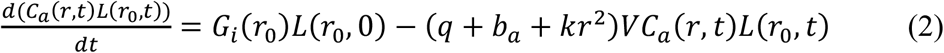

Where *L*(*r*_0_, 0) is the concentration of virus in the droplets with initial radius *r*_0_; *q* is the ventilation rate of the room; *b*_*a*_ is the death rate of coronavirus in aerosol, *V* is the room volume, and *kr*^2^ quantifies the deposition rate of droplets with particle size *r* on the horizontal environmental surface. *G*_*i*_(*r*_0_) is the generation rate of droplets with radius *r*_0_ exhaled by the infector. Denoted the total amount of droplets generated by the infector per hour to be *N*_*d*_, and the droplet size distribution to be *f*(*r*_0_), then there is *G*_*i*_(*r*_0_) = *N*_*d*_*f*(*r*_0_). At steady state 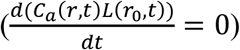, there is

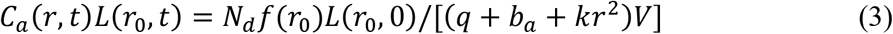

### Droplet route model

For droplet transmission route, there are mainly two ways: (1) inhalation of the virus carried in inspirable droplets with a diameter between 5 and 50 *μm* (Nicas and Jones, 2009; Lei et al., 2018). These inspirable droplets mainly deposited in the upper respiratory tract; (2) droplet spray of virus contained droplets onto the face membranes of susceptible individuals during talking and coughing. We assume that the trajectory of the droplets exhaled by the infector forms a jet. It is assumed that when coughing/talking, the cone angle formed by the jet is *α*, the mucosal area of a person is *A*_*m*_, at a horizontal distance of *S* (*S*≤ 2 m) from the infector, the virus concentration in the droplet is:

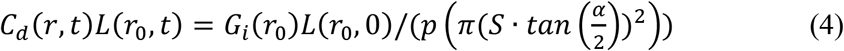

Assume that the exposure time of susceptible individuals in droplet transmission route is *T*_*d*_. Then the exposure dose caused by inhalation is:

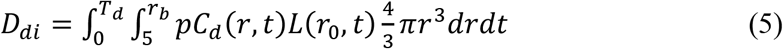

Where *r*_*b*_ is the maximum radius of inspirable droplets. *L*(*r*_0_, *t*) is the concentration of viable pathogens in the droplets with initial particle size *r*_0_ at the time t after being exhaled. For small droplets with particle size less than 10 μm, they have evaporated completely before being inhaled by the susceptible person, so *L*(*r*_0_, *t*) =0.25*L*(*r*_0_, 0), But for large droplets, they may not evaporate completely when being inhaled by the susceptible person, *L*(*r*_0_, *t*) is determined by its evaporation time and inhalation time of susceptible persons. Assume that Tm (s) is the traveling time (s) for the exhaled droplets from the source to reach a susceptible person a distance *S* away, *t*_*e*_(*r*_0_) is the evaporation time for the droplets with radius *r*_0_. There is

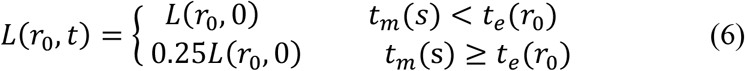

Xie et al. (2006) studied the evaporation time of droplets with different diameters, and a fitting function *t*_*e*_(*r*_0_)=7 × 10^−^*r*_0_^2^ was used in this study. *T*_*m*_(*s*) = *s*/*V*_*m*_, where *V*_*m*_ is droplet speed after exhalation. The exposure dose by spraying onto the mucous membrane is:

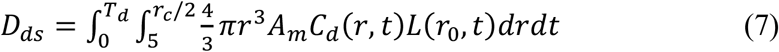

The average interpersonal distance between two individuals (*S*) during talking was set as 0.81 m (Zhang et al., 2020).

### Contact route model

By assuming 1% of virus contained droplets exhaled by the infector depositing on infector’s hands, the rate of virus deposition on the infector’s hand *S*_*s*_ is:

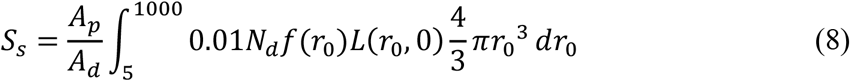

Where *A*_*p*_ and *A*_*d*_ represent the area of infector (adult) ‘s hand and large particle size droplet deposition area, respectively. With the deposition of viruses on hands, viruses on hands also lose activation and removed by hand hygiene. Denoted the natural inactivation rate of the virus on hands to be *η*_*np*_. And the hand hygiene frequency was assumed to be *c*_*hy*_ and the hand hygiene efficiency was assumed to be *e*_*hy*_. Hand hygiene (*e*_*hy*_) was considered to remove 90% of virus (Temime et al., 2009). In order to represent the discrete nurse hand hygiene process in the continuous governing ordinary differential equations, we made the following translation (Lei et al., 2020). After each hand hygiene, only a fraction (1–*e*_*hy*_) of viruses remains on hands. Hand hygiene occurs *c*_*hy*_ times per hour, and the time-average rate of pathogen removal due to hand hygiene is denoted by *η*_*hy*_. On average, there is 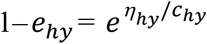, then *η*_*hy*_ = −log(1–90%)*c*_*hy*_.

At the steady state, the amount of virus on the patient’s hands 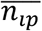 is

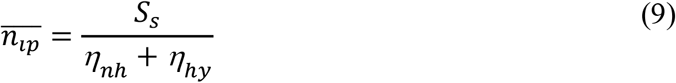

Where *η*_*nh*_ is the natural inactivation rate of the virus on hands.

Assume the infector and the susceptible individuals (adult) touch the environmental surface with rate *c*_*p1,s*_, the children touch the environmental surface with rate *c*_*p*2,*s*_, the frequency of hand-to-hand contact between the infector and the susceptible adult is *c*_*p1,p*2_, and between the infector and the susceptible children is *c*_*p1,p1*_. Then the amount of virus on the *i*th susceptible individual’s hand, *n*_*pi*_(*t*) (*i*=1 for adults, *i*=2 for young), and the *k*th environmental surface, *n*_*sk*_(*t*) (*k*=1 represents porous surface, *k*=2 represents non porous surface) could be calculated by equation (10-12)::

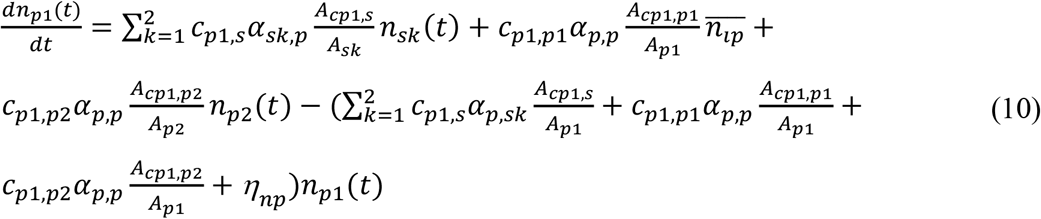

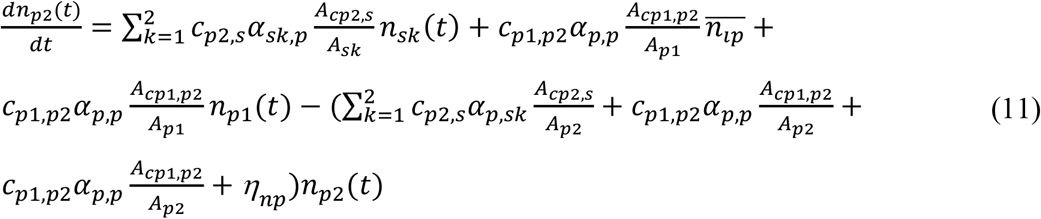

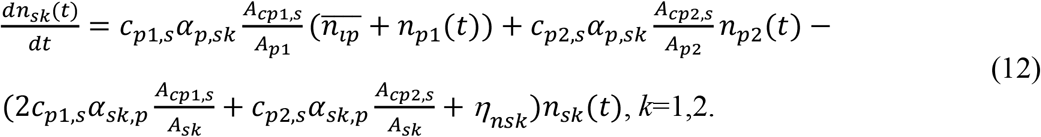

Where *α*_*p,sk*_,*α*_*sk,p*_ and *α*_*p,p*_ represent the transmission efficiency of the virus from hand to the *k*th environmental surface, from the *k*th surface to hand, and from hand to hand, respectively. *A*_*pi*_, *A*_*sk*_ *A*_*cpi,s*_ and *A*_*cp1,pi*_ represents the area of the *i* th susceptible individual’s hand, the area of the *k*th environmental surface, the contact area during hand-to-environmental surface contact, and the contact area during hand-to-hand contact, respectively. *η*_*nsk*_ is the inactivation rate of the virus on the *k*th environmental surface. Assumed that hand washing and surface cleaning are carried out at a certain frequency at some time points. And the hand washing frequency is assumed to be *r*_*p*_ with efficiency at *η*_*p*_. The surface cleaning frequency is *r*_*s*_, with efficiency at *η*_*s*_.

Then the total exposure dose of *i* th susceptible individual via contact route at the exposure time *T* can be calculated by equation (13):

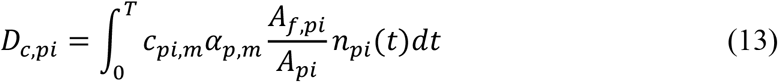

Where *c*_*pi,m*_ is the contact rate on the mucous membrane of *i*th susceptible individual, *α*_*p,m*_ is the transmission efficiency of the virus from hand to mucous membranes, *A*_*f,pi*_ is the contact area between hands and mucous membranes of the *i* th susceptible individual.

### Infection risk assessment

The negative exponential dose-response model (Lei et al., 2018) was used to estimate the infection risk, which implies that a single particle can start an infection, all single particles are independent of each other. The infection risk of individual during 1-day exposure can be calculated according to the following equation:

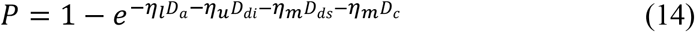

Where *η*_*l*_, *η*_*u*_, and *η*_*m*_ are the dose-response rate in low respiratory tract, upper respiratory tract and on mucous membranes, respectively. To our best knowledge, a dose-response relationship for both SARS-CoV-2 ancestral and Omicron variant as a cause of COVID-19 has not been reported. So in this study, we simply assumed that the dose-response rates for ancestral and Omicron variant were the same. Animal experiment suggested that airborne transmission was more efficient than fomite transmission (Port et al., 2021). And recent findings on SARS-CoV-2 also revealed that the angiotensin-converting enzyme 2, which was used to infect humans, was expressed at high levels in nose and low levels in the lower respiratory tract (approximately 4:1) (Hou et al., 2020). Thus, there is *η*_*l*_ ≥ *η*_*m*_. In this study, we suppose that *η*_*l*_: *η*_*u*_: *η*_*m*_ = 100: 1: 1 (Mizukoshi et al., 2021), though the 100:1 for infectivity in lower respiratory tract and upper respiratory tract/mucous membranes may be an overestimate, we set 100: 1 ratio mainly based on the data on influenza virus in humans, that for influenza, there is *η*_*l*_: *η*_*m*_ = 1000: 1 (Couch and Kasel, 1983; Nicas and Jones, 2009). For adults, *η*_*m*_ = 2.46 × 10^−3^, which was the dose-response rate for SARS-CoV, and was also the best available model in present study (Mizukoshi et al., 2021). For children, the dose response rates were assumed to be half of the these for adults, since studies reported that the susceptibility to SARS-CoV-2 of increases with age (Zhang et al., 2020; Lei et al., 2021). Detailed model parameterization was summarized in Supplementary Information Part 1 Model Parameterization.

### Sensitivity analysis

Some parameters had uncertainties that could not be controlled by the scenario settings. Thus we performed the following sensitivity analysis of these uncertain parameters (Supplementary Information Part 2 Sensitivity Analysis).

1. To assess the impact of hand-to-surface contact rates on the results, the hand-to-surface contact frequency at 2/h and 4/h were set respectively, in the baseline, the hand-to-surface contact frequency was 3/h.
2. To assess the impact of hygiene frequency on the results, the hand hygiene frequency at 0.25/h and 0.75/h were used, in the baseline, the hand hygiene frequency was 0.5/h.
3. To assess the impact of dose response rates on the results, different dose response rate for Omicron variant were set. In the baseline, for SARS-CoV-2 Omicron variant, the dose response rates in low respiratory tract, upper respiratory tract and on mucous membranes, were *η*_*m*_ = *η*_*u*_ = 2.46 × 10^−3^, *η*_*l*_ = 2.46 × 10^−1^, same to the ancestral strain. Since Omicron variant has higher infectivity than ancestral strain, in the sensitivity analysis, *η*_*m*_ = *η*_*u*_ = 5 × 10^−3^, *η*_*l*_ = 5 × 10^−1^, and *η*_*m*_ = *η*_*u*_ = 1 × 10^−2^, *η*_*l*_ = 1 were set respectively.
4. To assess the impact of virus deposition on different surfaces on the results, during asymptomatic or presymptomatic infection, the proportion of virus exhaled by infector deposited on porous were set as 69.5% and 29.5% respectively, thus the proportion of virus deposited on non-porous surfaces were set as 29.5% and 69.5% respectively. In the baseline, during asymptomatic or presymptomatic infection, we assumed that 49.5% and 49.5% of the virus contained droplets exhaled by the infector were partitioned to porous and non-porous surfaces.

## Results

### Estimation of the infection risk

The risks of each pathway and the overall risk depended on the virus concentration in saliva (Figure 2). For ancestral and Omicron strain, under same conditions, the overall infection risk was close, a little higher for Omicron strain (Figure 2). This was mainly due to the assumption that ancestral and Omicron strain had same dose response rates. But Omicron variant would have higher dose response rates since Omicron variant had higher infectivity. With higher dose response rates, the infection risk of Omicron variant would be higher. For both the susceptible child and the adult, when the virus concentration in saliva was 10^4^-10^8^ mRNA copies/mL, the overall infection risk of the susceptible child was about half that of the susceptible adult, since the dose response rate of the child was set as half of that of the adult. During asymptomatic or presymptomatic infection, for the susceptible child, the overall infection risk was 2.2×10^−4^ for virus concentration in saliva of 10^5^/mL, 2.2×10^−2^ for virus concentration in saliva of 10^7^/mL, and 0.89 for virus concentration in saliva of 10^9^/mL. For the susceptible adult, the overall infection risk was 4.4×10^−4^ for virus concentration in saliva of 10^5^/mL, 4.3×10^−2^ for virus concentration in saliva of 10^7^/mL, and 0.99 for virus concentration in saliva of 10^9^/mL. During symptomatic infection, for the susceptible child, the overall infection risk was 2.3×10^−4^ for virus concentration in saliva of 10^4^/mL, 2.3×10^−2^ for virus concentration in saliva of 10^6^/mL, and 0.90 for virus concentration in saliva of 10^8^/mL. For the susceptible adult, the overall infection risk was 4.7 × 10^−4^ for virus concentration in saliva of 10^4^/mL, 4.6 × 10^−2^ for virus concentration in saliva of 10^6^/mL, and 0.99 for virus concentration in saliva of 10^8^/mL. When the infector was symptomatic, the infector produced much more droplets, since cough could produce much more droplets than breathing and speaking, but the virus concentration in the saliva was supposed to be lower. Thus the overall infection risk of two susceptible individuals by asymptomatic or presymptomatic infection was close to that by symptomatic infection. This suggested that asymptomatic or presymptomatic infection could contribute about 50% of the SARS-CoV-2 transmission.

**Figure 2.**
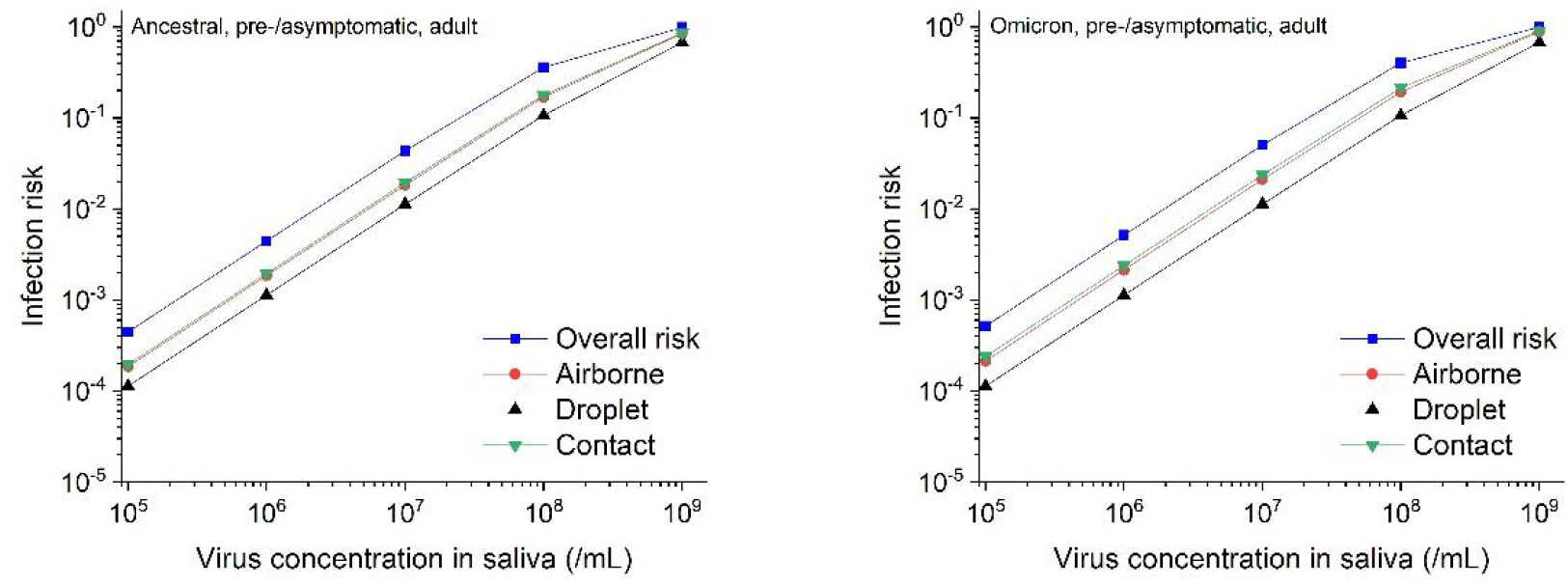
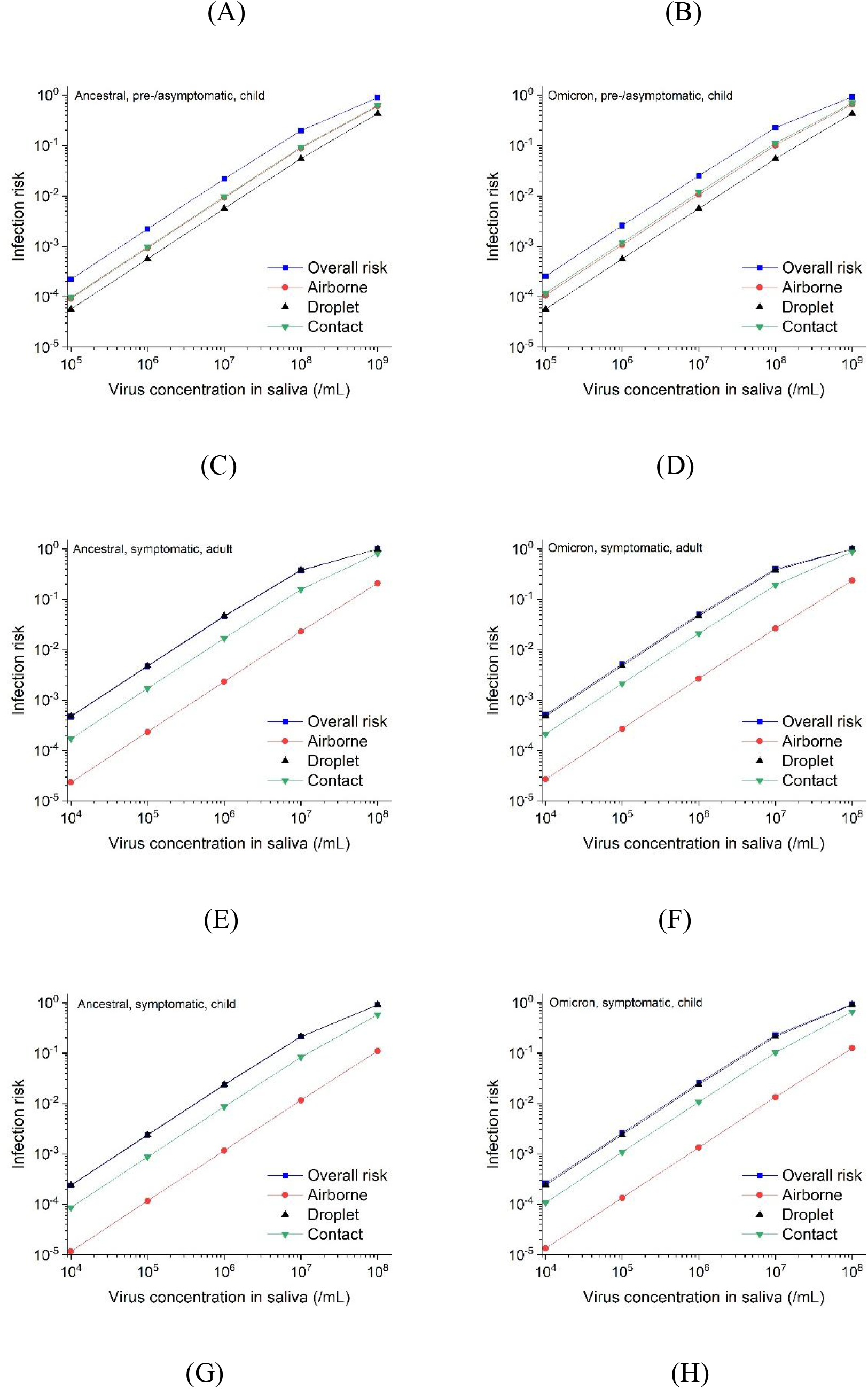
Absolute infection risk from each pathway and overall risk. (A, B, C, D) during asymptomatic or presymptomatic infection, with virus concentration in saliva at 10^5^-10^9^ RNA copies/mL respectively; (E, F, G, H) during symptomatic infection, with virus concentration in saliva at 10^4^-10^8^ RNA copies/mL respectively; (A, C, E, G) for ancestral strain; (B, D, F, H) for Omicron variant; (A, B, E, F) for the susceptible adult; (C, D, G, H) for the susceptible child.

The contribution of each pathway according to different virus concentration in saliva are shown in Figure 3. For both strains, the dominant routes during asymptomatic or presymptomatic were different to that during symptomatic infection. When the infector was asymptomatic or presymptomatic, all three routes contributed a role in SARS-CoV-2 transmission (Figure 3A, 3B). Contact route contribute most (37%-45%), followed by airborne route (34%-38%) and droplet route (21%-28%). When the infector was symptomatic, droplet route was the dominant pathway (48%-71%), followed by contact route (25%-42%). Airborne route played a negligible role (<10%). Compared with ancestral strain, though the role of contact route increased in Omicron variant transmission, the increase was slight (Figure 3).

**Figure 3.**
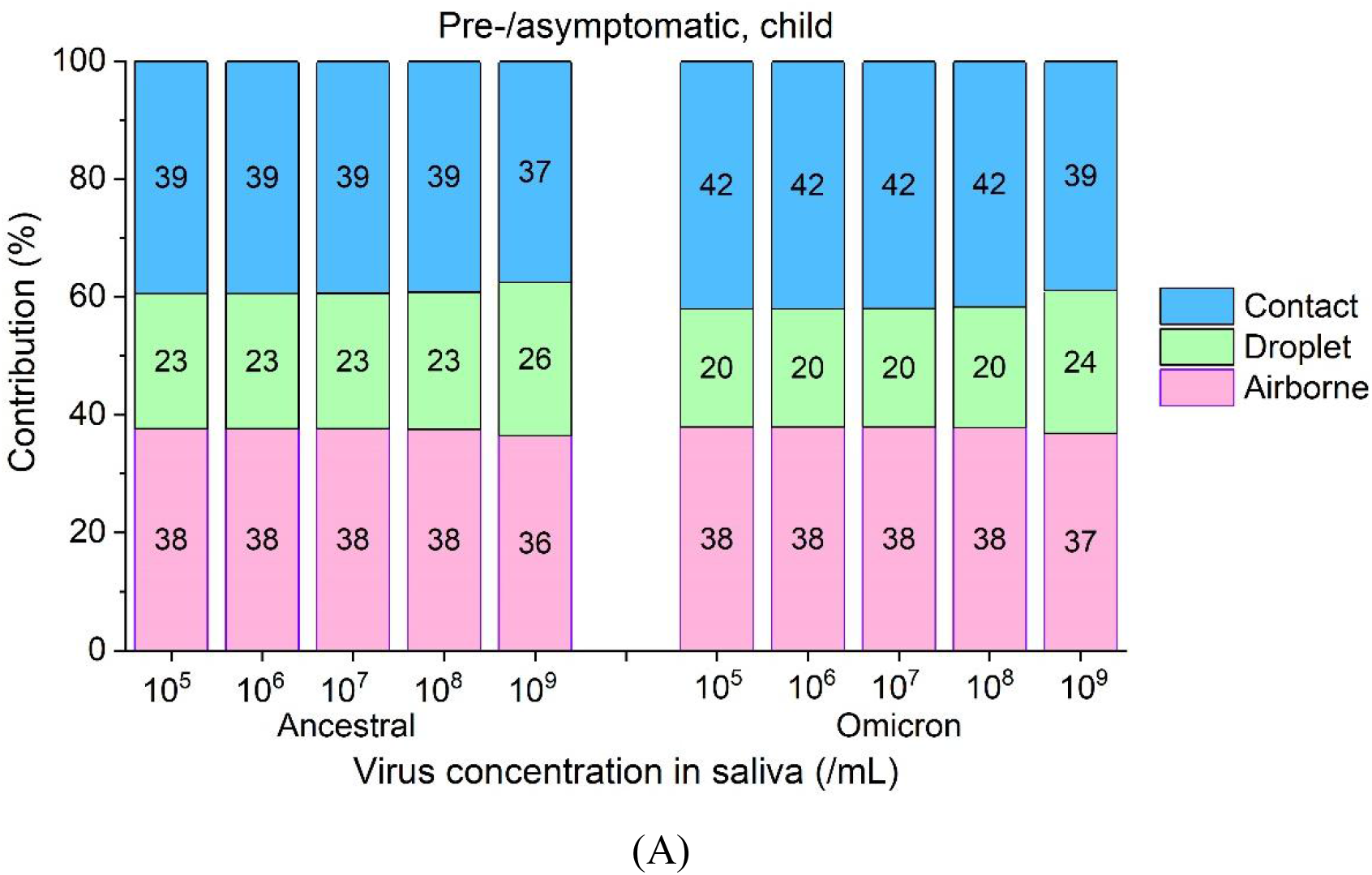
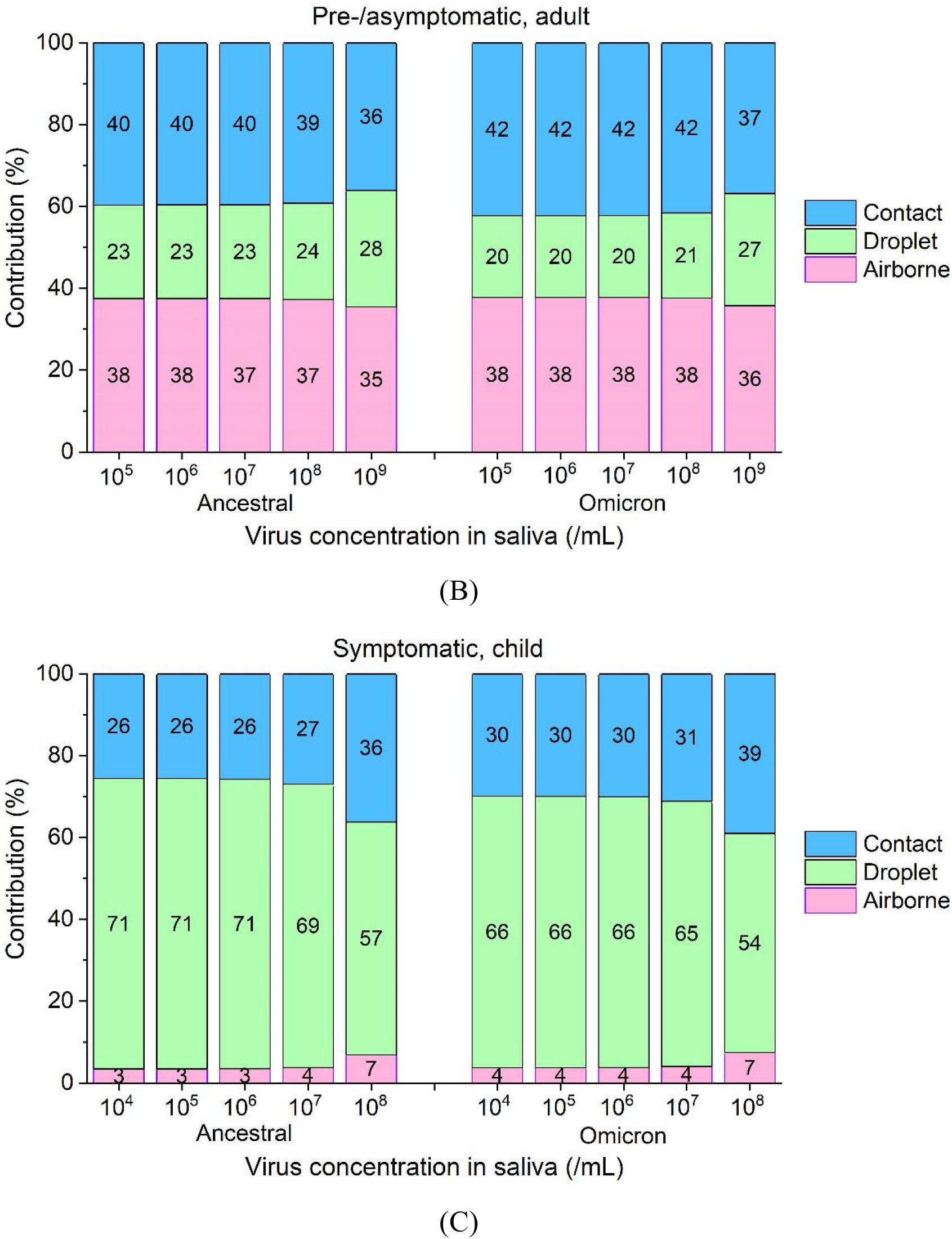
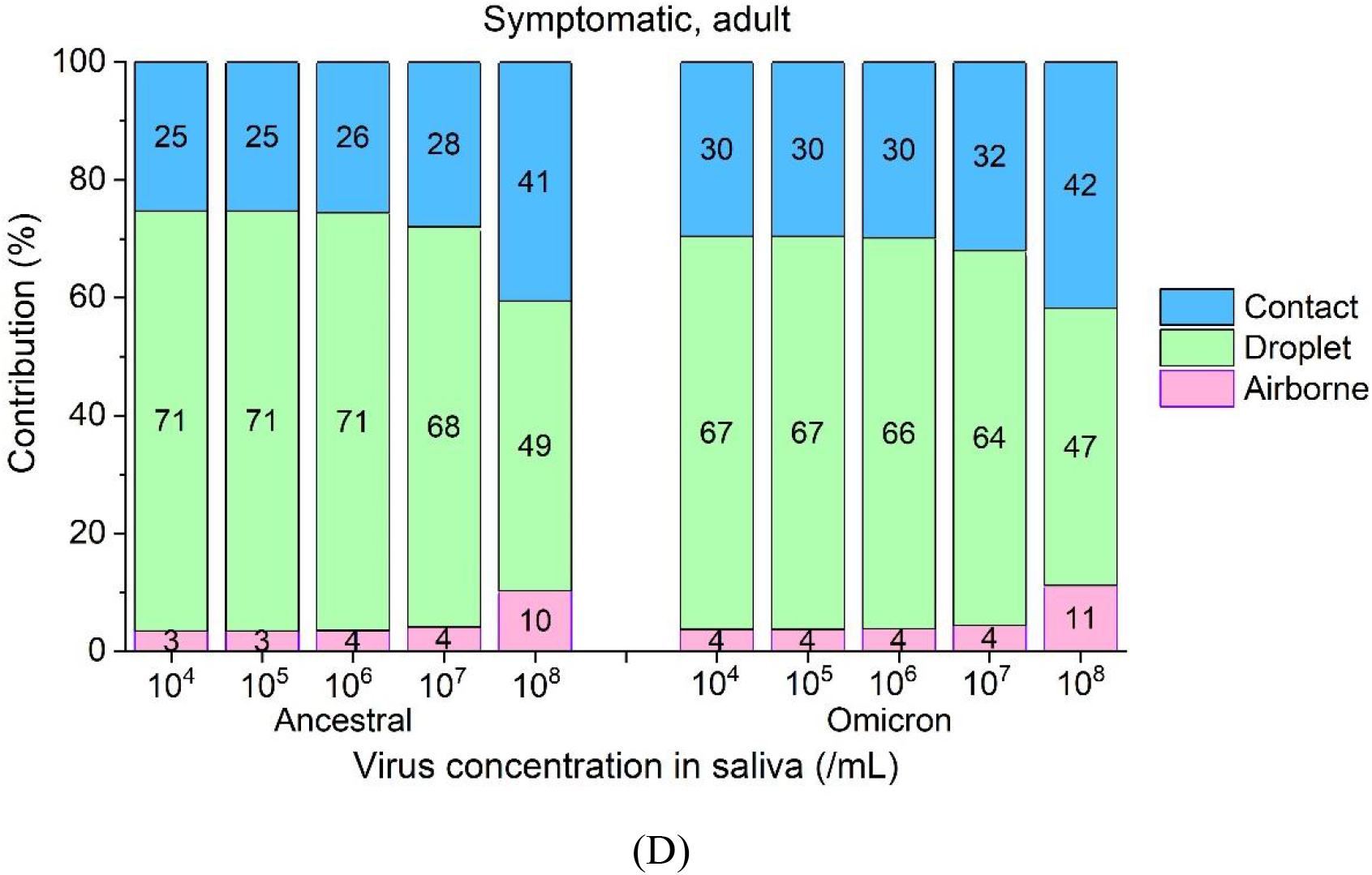
Contribution of airborne, droplet, and contact transmission routes to the (A, C) susceptible child and (B, D) the susceptible adult, (A, B) during symptomatic infection and (C, D) during asymptomatic or presymptomatic infection, for different saliva virus concentrations. Left: ancestral strain; Right: Omicron strain.

We also explored the relative contributions of direct and indirect contact routes in contact transmission of SARS-CoV-2 (Figure 4). In contact transmission of SARS-CoV-2 in household, indirect contact route dominated. This may be mainly due to that the frequency of hand-to-environmental surfaces contact (3 times per hour) was much higher than the frequency of hand-to-hand contact (0.1-0.5 times per hour), and the hand hygiene frequency was 0.5 per hour, but the environmental surfaces were not cleaned, so the virus concentration on environmental surfaces was higher than these on the hands (Supplementary Information Part 3 Figure S5). And because of the relative higher hand-to-hand contact between the susceptible child and the infector, thus direct contact route contributed more in contact transmission of SARS-CoV-2 for the susceptible child (Figure 4).

**Figure 4.**
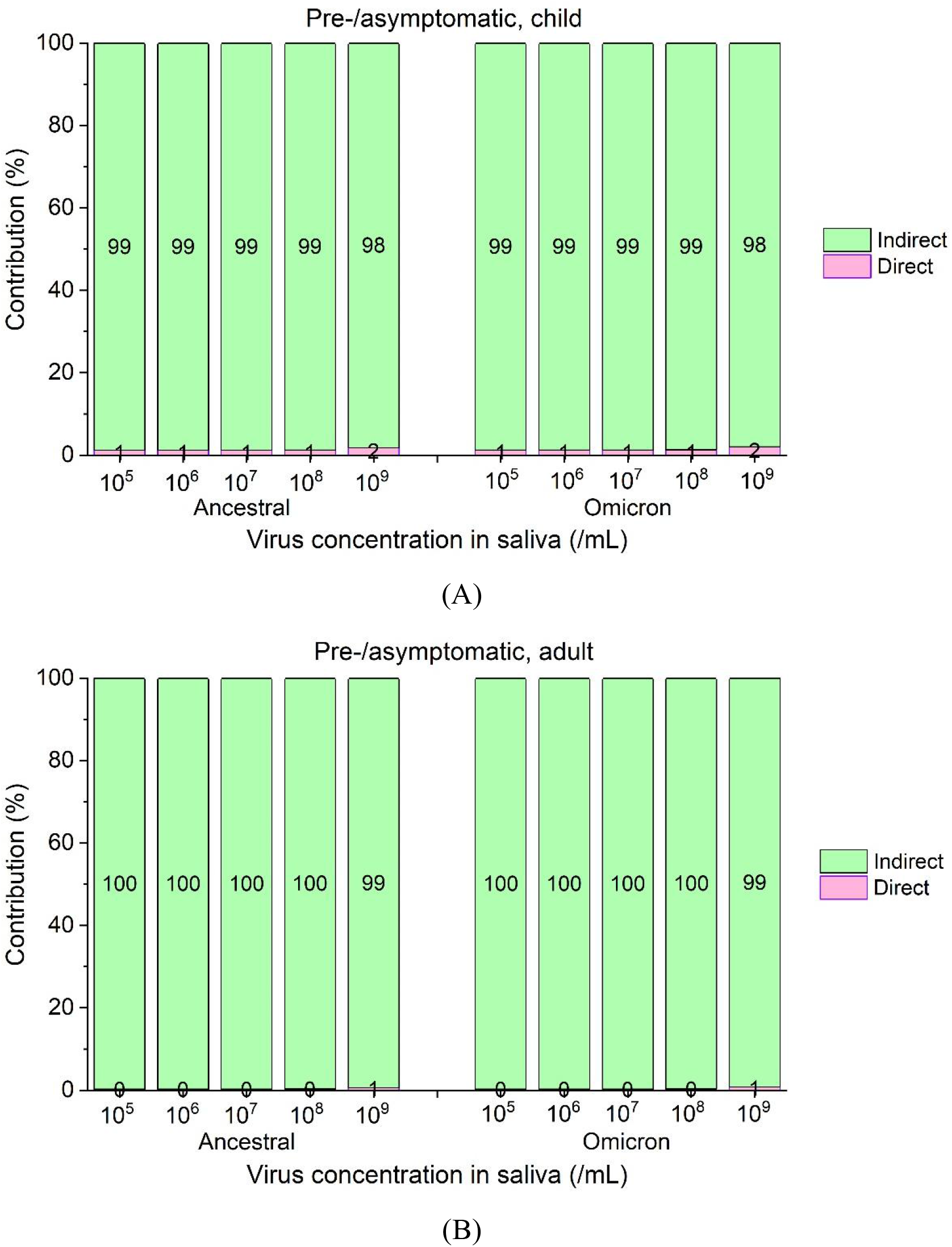
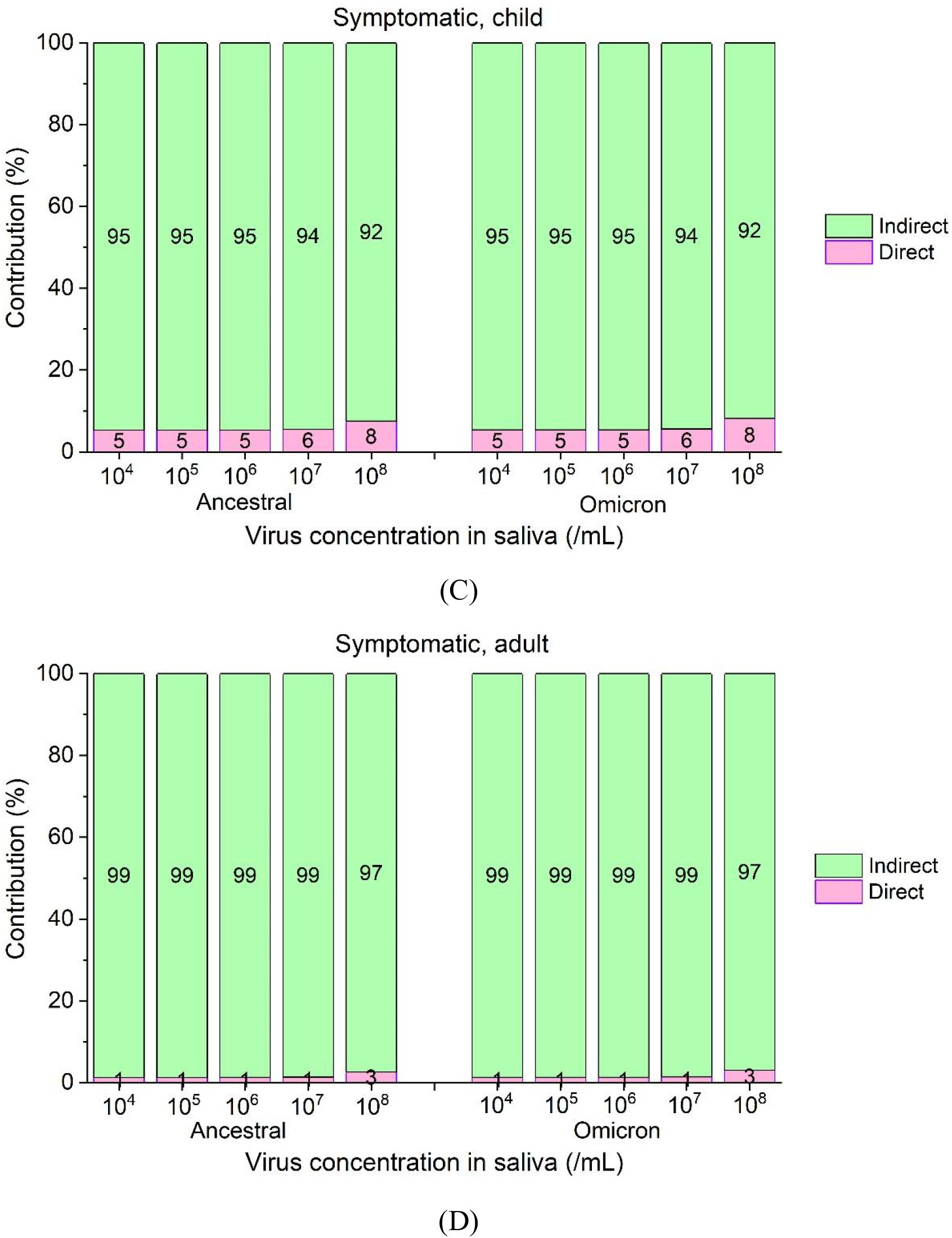
Contribution of direct and indirect (fomite) contact route in contact transmission of SARS-CoV-2. (A, C) susceptible child and (B, D) the susceptible adult, (A, B) during symptomatic infection and (C, D) during asymptomatic or presymptomatic infection, for different saliva virus concentrations. Left: ancestral strain; Right: Omicron strain.

## Discussion

The emergence of the new coronavirus variant strain Omicron has led to a rapid increase in the proportion of asymptomatic infections, making it more difficult to prevent and control the epidemic. In the scientific and effective control of human-to-human transmission of SARS-CoV-2, the relative importance of the pathways of exposure to SARS-CoV-2 have to be quantified, to determine the effective non-pharmaceutical interventions. In this study, based on the current knowledge about SARS-CoV-2 and its related information, we calculated the infection risk of SARS-CoV-2 from multiple pathways of exposure to SARS-CoV-2 in household settings and estimated the relative contribution of each pathway. We found that the relative importance of different routes during pre-/asymptomatic infection and symptomatic infection varied. In pre-/asymptomatic infection, all three routes contributed a role. In symptomatic infection, droplet route was the dominant pathway (48%-71%) and airborne route played a negligible role (contribution<10%). The main reason is that during pre-/asymptomatic infection, SARS-CoV-2 was mainly emitted via breathing and talking, and most droplets emitted from breathing were respirable droplets with diameter less than 10 microns. When the infector developed symptom, most viruses were emitted from coughing, since the volume of saliva from coughing was much larger than the volume of saliva from breathing and breathing. And droplets from coughing had relative larger size, so most exhaled droplets would deposit on environmental surfaces and inhale by susceptible individuals via droplet route. Concerned about the increase in the proportion of pre-/asymptomatic infections in Omicron strain (Garrett et al., 2022), the study of relative importance different routes in pre-/asymptomatic infection is very significant for SARS-CoV-2 prevention. In addition, for both pre-/asymptomatic and symptomatic infections, though SARS-CoV-2 Omicron variant was more stable on various environmental surfaces than ancestral strain, the contribution of contact route in SARS-CoV-2 transmission increased slightly for Omicron variant.

Previous studies had explored the relative contributions of different routes in SARS-CoV-2 ancestral variant transmission, via both animal model (Bao et al., 2020) and mathematical models (Jones, 2020; Mizukoshi et al., 2021). In the animal experiment study, the absolute risk of airborne, droplet and overall risk are 0, 3/10 and 7/13 respectively, thus simply considering inclusion and exclusion of contact transmission, the estimated contribution of airborne, droplet and contact route could be 0%, 57% and 43% respectively. This tendency was consistent with the results when the infector was symptomatic. In the modelling studies by Jones (2020) and Mizukoshi et al. (2021), they both considered the virus emission via coughing, and in the study by Jones (2020), the author even only considered the virus emission via coughing. Jones found that droplet and airborne routes predominated, contributing 35% and 57% respectively in hospital. Mizukoshi et al. (2021) found that airborne route was much less important than contact route and droplet route, only contributing 4%-10% in SARS-CoV-2 transmission in hospital. This was consistent with the results when the infector was symptomatic in this study. In addition, Mizukoshi et al. (2021) found that the role of droplet route decreased with the increase of virus concentration in saliva, contributing 65%-70% when the virus concentration was 10^1^-10^4^/mL, and 20%-46% when the virus concentration was 10^5^-10^8^/mL. When the virus concentration was low, the estimated contribution of contact route was consistent with the results in this study (67%-71%). But when the virus concentration was high, the estimated 20%-46% contributions were much lower than these in this study (48%-71%). The main reason could be that the estimated overall risk in the study by Mizukoshi et al. (2021) was much higher than our estimation under same virus concentration in saliva. When the infection risk by contact route was high, with further quick increase of virus concentration in saliva, the infection risk by contact route would increase slowly, while infection risk via other route would still increase quickly, thus the contribution of contact route decreased. The relatively low overall infection risk in this study was mainly due to the relatively small volume of droplets exhaled by talking (0.16 VS 3.2× 10^−4^ mL saliva per 100 second speaking) and coughing (4.4× 10^−2^ VS 8.1× 10^−3^ mL per cough) in this study.

SARS-CoV-2 transmission by fomites was demonstrated in animal model studies (Bao et al., 2020; Sia et al., 2020; Port et al., 2021). And the presence of SARS-CoV-2 has been reported on various surfaces (Guo et al., 2020; Mondelli et al., 2020). In the long-term care facilities with COVID-19 patients, the SARS-CoV-2 concentrations on environmental surfaces ranged from 1.3 to 3661.2 genomes/cm^2^, with median value at 76.6 genomes/cm^2^ (Dumont-Leblond et al., 2021). The estimated virus concentration on environmental surfaces in this study was close to the results from field measurements. The transmission of SARS-CoV-2 Omicron via packaging had been attracted widespread attention. Due to the virus inactivation of Omicron variant is lower than ancestral strain (Chin et al., 2022), we are beginning to worry about whether the risk of contact transmission is significantly increased. Since both Omicron and ancestral variant survival well on environmental surfaces, and hand hygiene played a key role in contact transmission of SARS-CoV-2. In our study, we found that the role of contact route increased slightly in Omicron variant transmission, no more than 5%. Therefore, contact transmission is not enough to cause too much concern. In contrast, we found that with the increase of pre-/asymptomatic infections in Omicron strain transmission (Garrett et al., 2022) compared with ancestral strain, airborne route should gain more attention in SARS-CoV-2 transmission and prevention. When the infector was symptomatic, airborne route played a negligible role (<10%), but this pathway became important (34%-38%) when the infector was asymptomatic or presymptomatic.

This study has several limitations. Firstly, the results depend on the model assumptions. For example, the relative contribution of droplet route was dependent on the emission of virus in respirable droplets and the exposure time of susceptible individuals. The hand hygiene frequency and the hand hygiene efficiency affected the results related to contact route. These parameters remain highly uncertain. To improve the accuracy of the model, it is desirable to update the data pertaining to SARS-CoV-2 in future studies. Secondly, it should be noted that the scenario we assume at home is relatively simple, we only considered the close contact distance of 0.81 m between two individuals, and contacts of infected and susceptible are only considered for 30 minutes of conversation.

In reality, there will be more complex activities, such as dining together, that may increase the risk of droplet and contact route. In other public places such as shopping malls, restaurants, etc., the dominant route may be different, thus we would further study scenarios of the other settings in the future study. Last, the behavioral settings of the susceptible adult and the susceptible child are the same, however, children may touch more surfaces and wash their hands less often, which may lead to a higher risk of contact transmission in children. Therefore, the overall infection risk of the susceptible child was about half that of the susceptible adult, this conclusion may vary under different settings. In a word, the overall risk of COVID-19 infection predicted by the model should be interpreted with caution, though within result comparisons remain informative.

## Data Availability

All data produced in the present study are available upon reasonable request to the authors

